# High gamma activity distinguishes frontal cognitive control regions from adjacent cortical networks

**DOI:** 10.1101/2021.08.13.21261980

**Authors:** Moataz Assem, Michael G. Hart, Pedro Coelho, Rafael Romero-Garcia, Alexa McDonald, Emma Woodberry, Robert C. Morris, Stephen J. Price, John Suckling, Thomas Santarius, John Duncan, Yaara Erez

**Affiliations:** Medical Research Council, Cognition and Brain Sciences Unit, University of Cambridge; Department of Neurosurgery, Cambridge University Hospitals NHS Foundation Trust; St George’s, University of London & St George’s University Hospitals NHS Foundation Trust, Institute of Molecular and Clinical Sciences; Neurophys Limited; Department of Psychiatry, University of Cambridge; Department of Medical Physiology and Biophysics, Instituto de Biomedicina de Sevilla (IBiS) HUVR/CSIC/Universidad de Sevilla/CIBERSAM, ISCIII, Sevilla, Spain; Department of Neuropsychology, Cambridge University Hospitals NHS Foundation Trust; Division of Neurosurgery, Department of Clinical Neurosciences, University of Cambridge; Behavioural and Clinical Neuroscience Institute, University of Cambridge; Cambridge and Peterborough NHS Foundation Trust; Department of Physiology, Development and Neuroscience, University of Cambridge; Department of Experimental Psychology, University of Oxford; Faculty of Engineering, Bar-Ilan University, Ramat-Gan, Israel; Gonda Multidisciplinary Brain Research Center, Bar-Ilan University, Ramat-Gan, Israel

## Abstract

Though the lateral frontal cortex is broadly implicated in cognitive control, functional MRI (fMRI) studies suggest fine-grained distinctions within this region. To examine this question electrophysiologically, we placed electrodes on the lateral frontal cortex in patients undergoing awake craniotomy for tumor resection. Patients performed verbal tasks with a manipulation of attentional switching, a canonical control demand. Power in the high gamma range (70-250 Hz) distinguished electrodes based on their location within a high-resolution fMRI network parcellation of the frontal lobe. Electrodes within the canonical fronto-parietal control network showed increased power in the switching condition, a result absent in electrodes within default mode, language, cingulo-opercular and somato-motor networks. High gamma results contrasted with spatially distributed power decreases in the beta range (12-30 Hz). These results confirm the importance of fine-scale functional distinctions within the human frontal lobe, and pave the way for increased precision of functional mapping in tumor surgeries.

## Introduction

How the frontal cortex is anatomically and functionally organized to control cognition remains puzzling. Data from functional magnetic resonance imaging (fMRI) increasingly suggest that the lateral frontal cortex of the human brain is divided into functionally separate regions (Glasser et al., 2016). Temporal correlations in resting-state fMRI data cluster these regions to different whole-brain networks. Using the high-resolution multi-modal cortical parcellation of the human connectome project (HCP), for example, a recent study suggests lateral frontal participation in at least five functional networks: fronto-parietal (FPN), default-mode, cingulo-opercular, language and sensori-motor networks (Ji et al., 2019). These functional networks contrast with traditional coarser clinical divisions, e.g., dorso-lateral, ventro-lateral, orbito-medial, fronto-polar (Stuss, 2011). But a related electrophysiological signature of frontal divisions remains elusive. For example, the non-invasive electro/magneto-encephalography (EEG/MEG), remain blind to fine-grained frontal territories owing to their limited spatial resolution (Farahibozorg et al., 2018).

Here we use electrocorticography (ECoG) and fMRI fine-scale distinctions of the lateral frontal lobe to examine functional distinctions in electrodes placed directly on the frontal cortical surface during awake neurosurgery. ECoG offers high spatial resolution access to local field potentials (LFPs) in the gamma frequency range (30-250 Hz), a well-established index of local cortical processing that cannot be detected with non-invasive methods (Crone et al., 2006; Lachaux et al., 2012). Our focus is on the FPN, and its role in executive or cognitive control (Duncan, 2010; Cole et al., 2013). Cognitive control tasks have consistently highlighted increased fMRI activations in the FPN with increased cognitive demands (Fedorenko et al., 2013; Assem et al., 2020; Shashidhara et al., 2020). In ECoG studies, power increases in the gamma range have been linked to heightened attentive states (Bouyer et al., 1981), processing of attended stimuli (Fries, 2001; Ray et al., 2008; Szczepanski et al., 2014; Helfrich and Knight, 2016), working memory (Howard, 2003; Mainy et al., 2007; Haller et al., 2018), response inhibition (Swann et al., 2012; Fonken et al., 2016) and responses to abstract rules (Voytek et al., 2015). While the link between cognitive control and gamma increases in the frontal lobe is well documented, whether gamma activity is related to the functional divisions revealed by fMRI remains unclear. This is because most ECoG studies broadly label electrodes according to cortical gyri and sulci. However, cortical curvature in association areas is a poor predictor of the location of functional areas (Coalson et al., 2018) leaving the link between gamma increases and frontal networks unknown.

Using two independent fMRI-based frontal parcellations, including the high-resolution HCP multi-modal MRI network parcellation (Ji et al., 2019), we assigned each electrode to its nearest resting-state fMRI network. To manipulate cognitive control demand, similar to fMRI manipulations that activate the FPN (Fedorenko et al., 2013; Assem et al., 2020), we used two tasks, simple counting (1 to 20; count) versus switching between counting and reciting the alphabet (1, a, 2, b, 3, c, up to 20; switch). Following a previous demonstration of the feasibility of ECoG for cognitive control mapping (Erez et al., 2021), here we use dense coverage of the frontal cortex to characterize the power modulations associated with increased cognitive demand. We predicted that increased high gamma responses in the switching compared to the simple counting condition would be localized specifically to regions of the FPN. Often, increases in high gamma activity are associated with decreases in beta band power. Though these two changes may be functionally linked (Helfrich and Knight, 2016), previous data suggest that beta decreases may be more widespread than gamma increases (Fellner et al., 2019). Accordingly, we predicted beta decreases both within and outside the FPN.

Our results demonstrate a distinct pattern of gamma activity within the FPN compared to other networks and confirm the importance of fine-scale functional distinctions within the human frontal lobe. Clinically, identifying the regions that support cognitive control intraoperatively using electrophysiology could provide valuable information to guide resection and prevent commonly-seen deficits following surgery (van Loon et al., 2015). Our results therefore pave the way for increased precision of functional mapping of the frontal lobe in tumor surgeries.

## Materials and Methods

### Patient recruitment

Twenty-one participants were recruited from a pool of patients with glioma undergoing awake craniotomy for tumor resection at the Department of Neurosurgery at Cambridge University Addenbrooke’s Hospital (Cambridge, UK). Data from thirteen patients were complete and were included in the study (age range 22-56; 6 males; see **Table 1** for patient demographics). Data from the remaining eight patients were excluded either due to technical difficulties (n=6; e.g. inability to localize electrodes correctly during surgery) or an inability to perform the tasks (n=2). All study procedures were approved by the East of England - Cambridge Central Research Ethics Committee (REC reference 16/EE/0151). All patients gave written informed consent to participate and were aware that the research would not benefit themselves, or impact their clinical care before, during or after surgery.

**Table 1.**
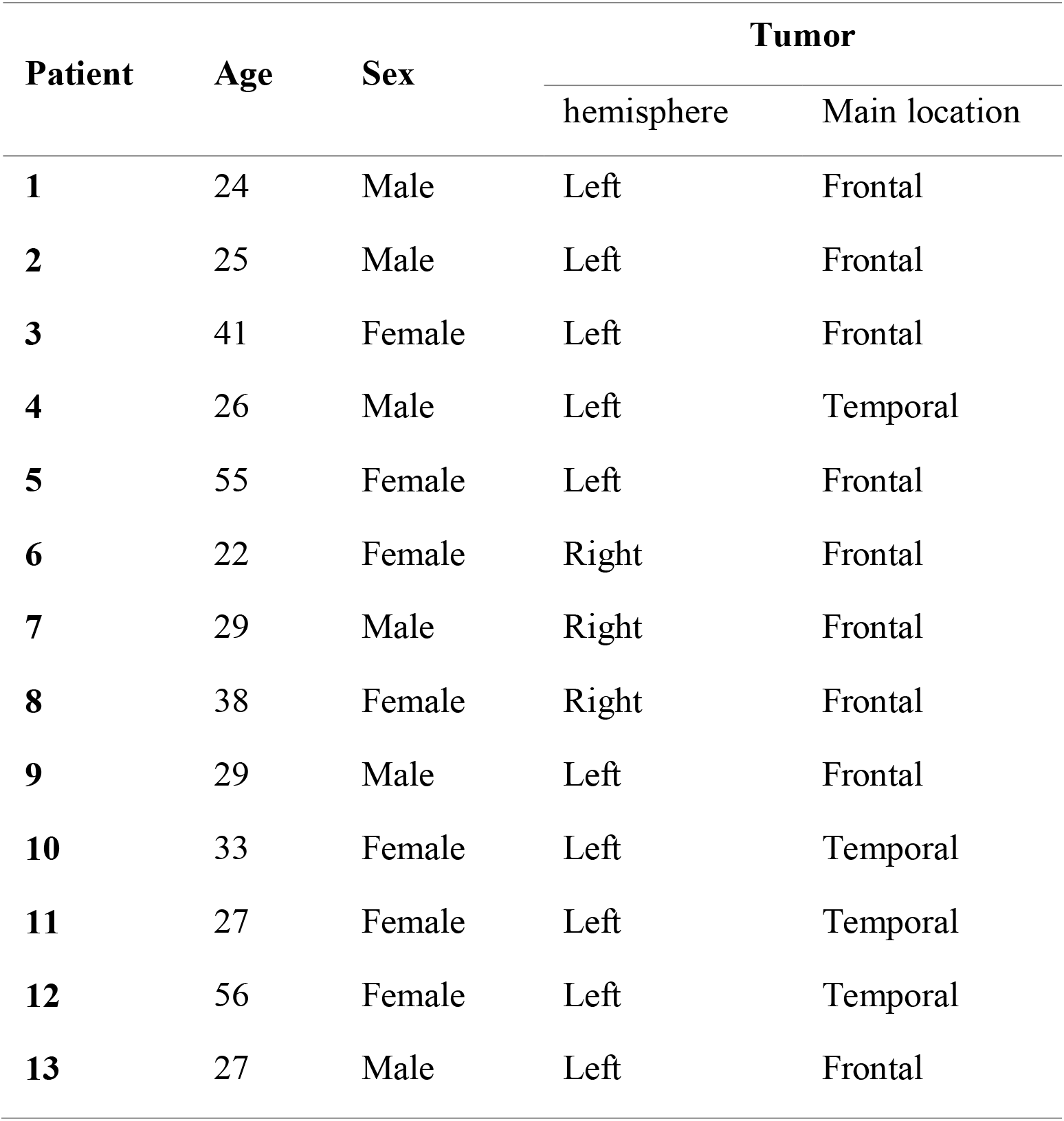
Patient demographics

### Experimental procedures

The patients were familiarized with the tasks during standard pre-operative clinic visits and as part of a pre-operative research-dedicated assessment. During the surgery and following the craniotomy, EEG (electroencephalogram) was recorded from scalp electrodes C3, C4 and Cz referenced to Fz for the purposes of monitoring anaesthetic depth and stimulation-related seizure activity. When the sedation was stopped the EEG evolved from high voltage, semi-rhythmic alpha and theta range components to a continuous trace consisting of the faster beta range frequencies (>13Hz) and low voltage activities in all derivations until the patients were awake. These low voltage beta range rhythms (normal awake activity) persisted during wakefulness and only then did cognitive testing commence. In all patients, testing was performed prior to tumor resection, except for one patient, where testing was performed after partial resection for clinical reasons. **Figures 1a, b** illustrate the intraoperative setup and cognitive tasks. During testing, all personnel in the surgical theatre were asked to limit their conversations to minimize disruptions. Patients performed one baseline task and two cognitive tasks. For the baseline task, the patients were asked to stay calm and remain silent for a period of 2-3 minutes (rest). The two cognitive tasks were simple counting (1 to 20; count) and alternation between counting and reciting the alphabet (1, a, 2, b, 3, c, up to 20; switch). Each task condition was repeated for 2-5 trials (median for both = 4 trials) based on each patient’s ability and time constraints during the surgery. Trial onset and offset markers were manually recorded on the acquisition system. Trial durations were 20.1±7.4s and 29.4±9.4s for the count and switch conditions, respectively. Most patients were instructed to alternate between trials of the count and switch conditions, though on a few occasions some trials of the same condition were performed in succession. Only correctly performed trials (i.e., no errors in simple or alternate counting) were included in the analysis (e.g., a failed switch trial that was excluded: 1, a, 2, b, 3, b, 4, b, 5, b, 6, b…).

**Figure 1.**
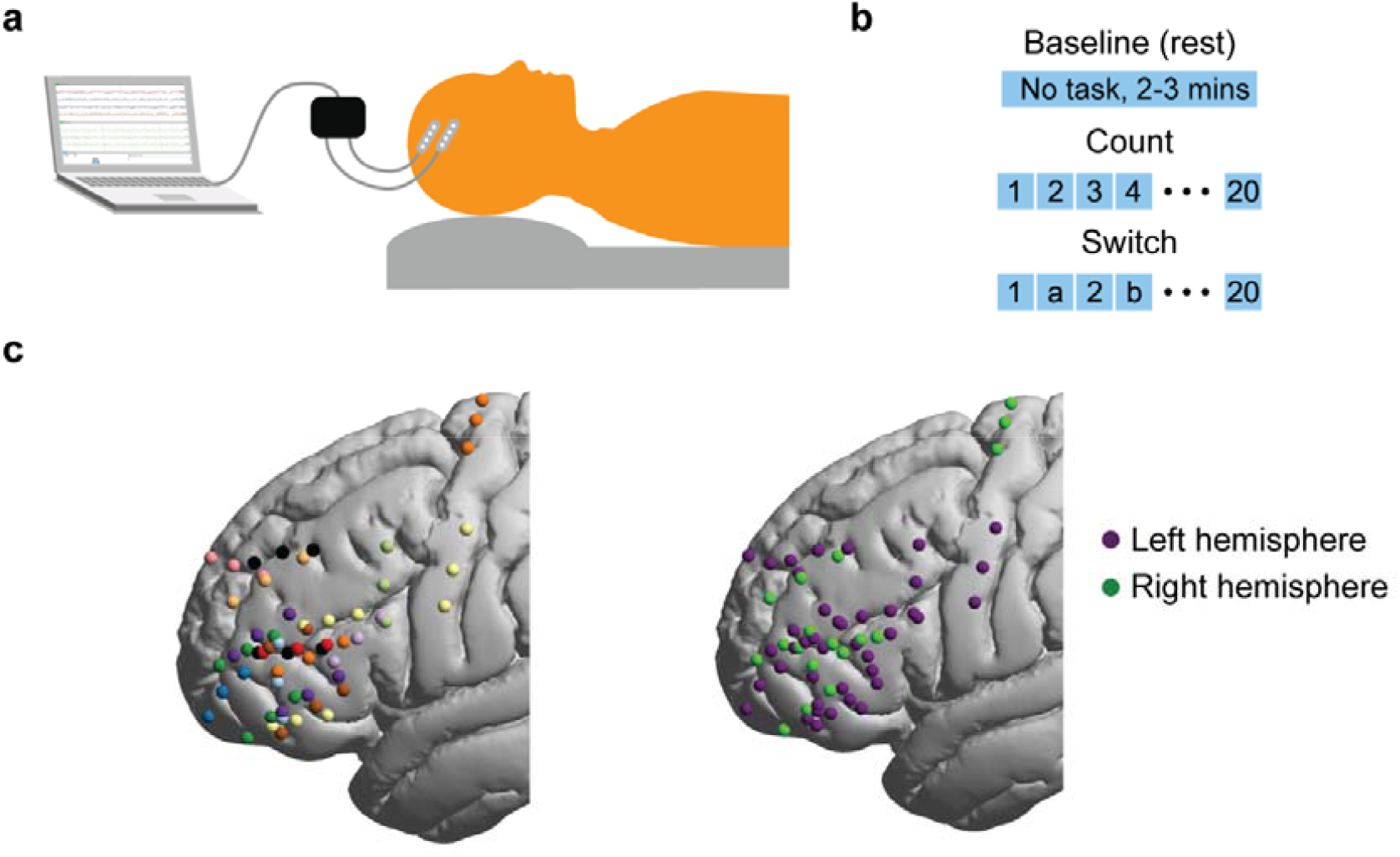
Intraoperative ECoG setup and electrode localization. (a) Intraoperative setup: Patient is awake during the three experimental conditions and the electrophysiological signals are simultaneously recorded using electrode strips placed directly on the cortical surface. (b) Experimental conditions: one rest (no task) and two verbal tasks. The count task involved simple counting from 1 to 20. The switch task involved alternating between numbers and letters. (c) *Left*: Electrode distribution for each patient in a separate color (13 patients, 59 electrodes after bipolar re-referencing). Electrodes from both hemispheres are projected onto the left. *Right*: hemispheric distribution of electrodes.

### MRI acquisition

MRI data were acquired pre-operatively using a Siemens Magnetom Prisma-fit 3 Tesla MRI scanner and 16-channel receive-only head coil (Siemens AG, Erlangen, Germany). Structural anatomic images were acquired using a T1-weighted (T1w) MPRAGE sequence (FOV = 256 mm x 240 mm x 176 mm; voxel size = 1 mm isotropic; repetition time (TR) = 2300 ms; echo time (TE) = 2.98 ms; flip angle = 9 degrees).

### Electrode localization

The extent of craniotomy of all patients was determined by clinical considerations to allow for tumor resection. Based on the craniotomy size and location, one to three electrode strips were placed on the cortical surface in regions judged by the neurosurgeon to be healthy (i.e. macroscopically not containing tumor). Strips placed on the tumor or outside of the frontal and motor cortices were excluded from analysis. Each strip was composed of four electrodes. Two types of strips were used with electrode diameter either 5 mm (MS04R-IP10X-0JH, Ad-Tech, Medical Instruments corporation, WI, USA) or 3 mm (CORTAC 2111-04-081, PMT Corporation, MN, USA). For both strip types, electrode spacing was 10 mm centre to centre.

Electrode locations were determined either using (1) an automated method with a probe linked to a stereotactic neuronavigation system (StealthStation® S7® System, Medtronic, Inc, 24 Louisville, CO, USA) or (2) a semi-manual grid method using intraoperative photographs and a grid-like delineation of cortical sulci and gyri. Most electrodes (51/79) were localized using the automated method, but due to occasional technical limitations, 28 electrodes were localized using the grid method. Both methods are detailed below.

#### (1) Stereotactic neuronavigation

A hand-held probe was placed at the centre of each electrode, automatically registering its physical coordinates, using the neuronavigation system, to the patient’s native high resolution preoperative T1w scan. In some cases, coordinate data were available for only two or three out of the four electrodes in each strip. This was due to either time constraints during the surgery or because an electrode was located underneath the skull, excluding probe placement. Each patient’s native T1w scan was linearly co-registered with the MNI template volume at 2 mm resolution using FLIRT as implemented in FSL (Jenkinson et al., 2012) using 12 degrees of freedom (full set affine transformation) and the correlation ratio cost function. The resulting native-to-MNI transformation matrix was then used to convert native electrode coordinates to MNI coordinates.

#### (2) The grid method

This follows the method described in (Havas et al., 2015) and (Ojemann et al., 1989). (a) Visible major sulci were delineated on the intraoperative photographs: precentral sulcus, sylvian fissure, inferior and superior frontal sulci. Spaces between these sulci were populated by vertical lines (1.5 cm apart) to create a grid-like structure. (b) A grid was created in the same way on a template cortical reconstruction of the MNI volumetric map (reconstructed using the HCP structural preprocessing pipeline 4.0.0; https://github.com/Washington-University/HCPpipelines). (c) MNI coordinates for each electrode were extracted by manually marking its approximate location on the template cortical grid while visualized using the Connectome Workbench v1.4.2 (https://www.humanconnectome.org/software/get-connectome-workbench). As the template cortical reconstruction is co-registered with its MNI volumetric version, it facilitated the automatic transformation of any point marked on the surface back to its MNI volumetric coordinates.

For both localization methods, electrode displacements due to brain shifts following the craniotomy were compensated for by back-projecting the electrode locations onto the cortical surface along the local norm vector (Hermes et al., 2010) as implemented in the fieldtrip (v20160629) protocol for human intracranial data (Stolk et al., 2018).

### Electrode network labelling

To relate electrode locations with frontal lobe parcellation identified by fMRI, we used two cortical parcellation schemes to ensure the robustness of the results: the Cole-Anticevic brain parcellation based on the high resolution data of the human connectome project (HCP) (Ji et al., 2019) and the canonical 7 network parcellation by (Yeo et al., 2011). For the surface based HCP-based parcellation, we first found the MNI coordinates associated with each surface vertex using the Connectome Workbench (v1.5) function wb_command –surface-coordinates-to-metric (using a group average midthickness surface as it provides the most accurate link between surface and MNI volumetric coordinates). For the Yeo et al parcellation, network labels were available in volumetric format (fieldtrip atlas) and needed no surface to volume transformation. Next, for each of the parcellation schemes, we related each electrode to its closest voxel coordinates (shortest Euclidean distance, pdist2 function in MATLAB v2018a) and assigned the electrode the most frequent network label of all voxels within a 5 mm distance.

### Electrophysiological data acquisition and analysis

Data were recorded using a 32-channel amplifier (Medtronic Xomed, Jacksonville, FS, USA) sampled at 10 KHz. Potential sources of electrical noise such as the surgical microscope, patient warming blanket, and IV pumps were identified and repositioned to avoid signal contamination. The data were recorded via dedicated channels on the acquisition system and two Butterworth online filters were applied: a high-pass filter at 1 Hz and a low-pass filter at 1500 Hz. A ground needle electrode was connected to the deltoid muscle and the electrodes were referenced to a mid-frontal (Fz) spiral scalp EEG electrode.

Data were analyzed offline using EEGLAB (v13.6.5b) and custom MATLAB scripts. The data were downsampled to 2 kHz then re-referenced using a bipolar scheme to detect any activity changes with the highest spatial resolution as well as to avoid contamination of high frequency signals by scalp muscle artefacts detected by the Fz electrode. The last electrode on the strip was excluded from analysis; i.e. for a four-electrode strip, electrode pairs 1-2, 2-3 and 3-4 were used and assigned to electrode positions 1, 2 and 3, respectively. The location of electrode 4 was discarded. Thus, out of the original 79 electrodes, re-referenced data from 59 were used for further analysis (**Figure 1c**). Out of these 59 electrodes, 41 were on the left hemisphere and 18 on the right. 25 electrodes were placed on the middle frontal gyrus (MFG), 28 on the inferior frontal gyrus (IFG) and 6 on motor cortex.

A notch filter was applied at 50 Hz and its harmonics to remove line noise. Notch filtering was also applied at 79 Hz and its harmonics to remove additional noise observed in the data, likely due to equipment in the surgical theatre.

The power spectral density (PSD) (Figure 3b) was estimated using pwelch in MATLAB (2018a) for each condition and each trial separately. Percentage of power change for each frequency was calculated as [(power in switch/power in count) – 1] * 100.

Data were then bandpass filtered into 6 classical frequency bands (delta: 1-4 Hz, theta: 4-8 Hz, alpha: 8-12 Hz, beta: 12-30 Hz, low gamma (LG): 30-70 Hz, high gamma (HG): 70-250 Hz). Instantaneous power of the timeseries was obtained by squaring the absolute amplitude envelope of the Hilbert transformed data.

The power timeseries data were then segmented into separate conditions and trials. Because trial onset and offset markers were manually recorded, 2s from the beginning and end of the rest trial and 1s from each task trial were excluded to account for human reaction time related error. For the switch trials, a further 3s from the beginning of each trial was excluded to discard the initial easy phase of this task (e.g., 1, a, 2, b, 3, c). One power value for each condition was obtained by concatenating its data across trials and averaging across time points. For each pair of conditions (switch>count, count>rest), the percentage of signal change was computed as: [(power in condition 1/power in condition 2) – 1] * 100.

For each electrode, a permutation testing approach was used to statistically compare power change across each pair of conditions. For each electrode, the instantaneous power timeseries of all task trials from both conditions were concatenated serially to form a loop in the same order in which they were conducted. To close the loop, the end of the last trial was joined to the beginning of the first trial. All trial onset/offset markers were then shifted using the same jitter (randomly generated for each permutation), allowing them to “rotate” along the data loop. This rotation approach was used to generate surrogate power data while preserving trial lengths and the temporal correlations in the data. After the rotation, we computed the mean power (for each condition) and power ratio (across conditions) based on the new trial markers. Applying this rotation approach on the timeseries of the power rather than the raw data ensured that there were no artefacts in the form of sudden power changes at the points of trials concatenation. This process was repeated 100,000 times to create a surrogate distribution against which two-tailed statistical significance was calculated (percentile ranks 97.5 and 2.5) for each electrode.

To statistically compare power changes between networks we used a linear mixed effects modelling approach with network identity as fixed effects and intercept grouped by subjects as random effect. This analysis was performed in MATLAB (2018a) using the fitlme function.

## Results

### Switch>count contrast reveals local increases in gamma power contrasting with widespread decreases in beta power

Across 59 electrodes (**Figure 1c**), we first examined spectral power changes with increased demand (switch>count contrast) for three classical frequency bands: high gamma (HG, 70-250 Hz), low gamma (LG, 30-70 Hz) and beta (12-30 Hz). In this contrast, positive values indicate power increases in switch compared to count, while negative values indicate power decreases. For the HG and LG bands, electrodes with significant power modulations showed predominantly increases (**Figure 2)**, and all but one of the electrodes that showed significant LG increases also showed HG increases. In contrast, changes in the beta band showed predominantly power decreases (**Figure 2)**. There was only partial overlap between locations of electrodes showing gamma increases and beta decreases (for HG, *r*=0.54, *p*<0.0001; for LG, *r*=0.32, *p*=0.003).

**Figure 2.**
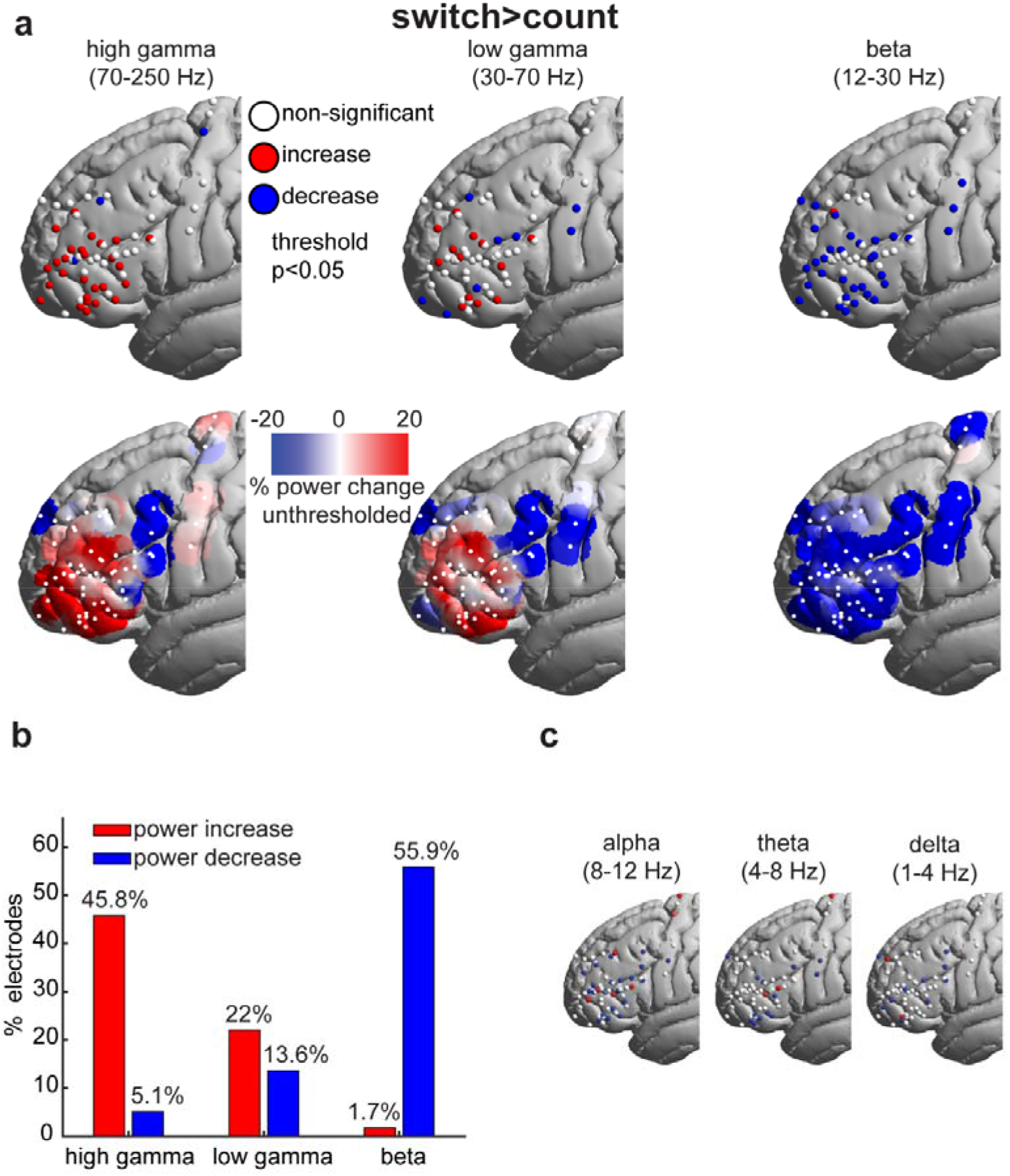
Switch>count spectral power modulations. (a) *Top*: Electrodes with significant (thresholded at *p*<0.05, uncorrected) power increases (red), decreases (blue) and non-significant changes (white). *Bottom*: Unthresholded average smoothed data. Power for each electrode (white dots; including electrodes with non-significant power changes) was spatially smoothed such that the value at each surface vertex is the average of the overlapping powers within a sphere of 10 mm radius. (b) Proportion of electrodes (out of a total of 59) showing significant power modulations for each of the frequency bands. (c) Electrodes colored as in the top row of (a) for the remaining three frequency bands.

Importantly, gamma power increases were spatially circumscribed, while beta decreases were more broadly distributed across the recording area. To quantify this observation, we compared the Euclidean distances between all pairs of electrodes with significant power changes for each of the three bands. We found that distances between significant electrodes for the HG (and to some extent the LG) bands were smaller than for the beta band (unpaired t-test; HG>beta: t_877_=-7.8, *p*=1.5×10^−14^; LG>beta: t_604_=-2.2, *p*=0.03; confirmed after excluding electrodes lying on the motor cortex: HG>beta: t_784_=-3.5, *p*=0.0005; LG>beta: t_511_=-0.98, *p*=0.33). Although this measure is, at least in part, affected by the spatial distribution of the electrodes, it nevertheless captures the distributed nature of the beta decreases compared to the more focal increases in the gamma range.

For completeness, we also examined power modulations in the lower frequency bands (delta, theta, alpha). These bands showed a similar picture to the beta band, with predominantly power decreases though with a more fragmented spatial arrangement (**Figure 2c**).

Taken together, these results show that, along the lateral frontal cortex, increased cognitive demand is associated with a localized increase in high frequency power and a spatially distributed decrease in low frequency power.

### High gamma power distinguishes fMRI-defined FPN from adjacent cortical networks

Next we asked how electrode power modulations compare to the canonically defined fMRI networks in the frontal lobe. We assigned each of 59 electrodes to its nearest resting-state fMRI network (**Figure 3a; see Materials and Methods**). 36 electrodes were located within the FPN, while 23 were outside this network [8 language (LANG), 5 default-mode (DMN), 4 cingulo-opercular (CON), 6 somato-motor (MOT)]. First we sought an overview of raw differences in PSDs between FPN and non-FPN electrodes. **Figure 3b** indeed suggests the major differences are broadly aligned with our beta, low gamma and high gamma divisions. Next we compared spectral power changes between networks for the switch>count contrast, focusing on the three classical frequency bands: high gamma (HG, 70-250 Hz), low gamma (LG, 30-70 Hz) and beta (12-30 Hz).

**Figure 3.**
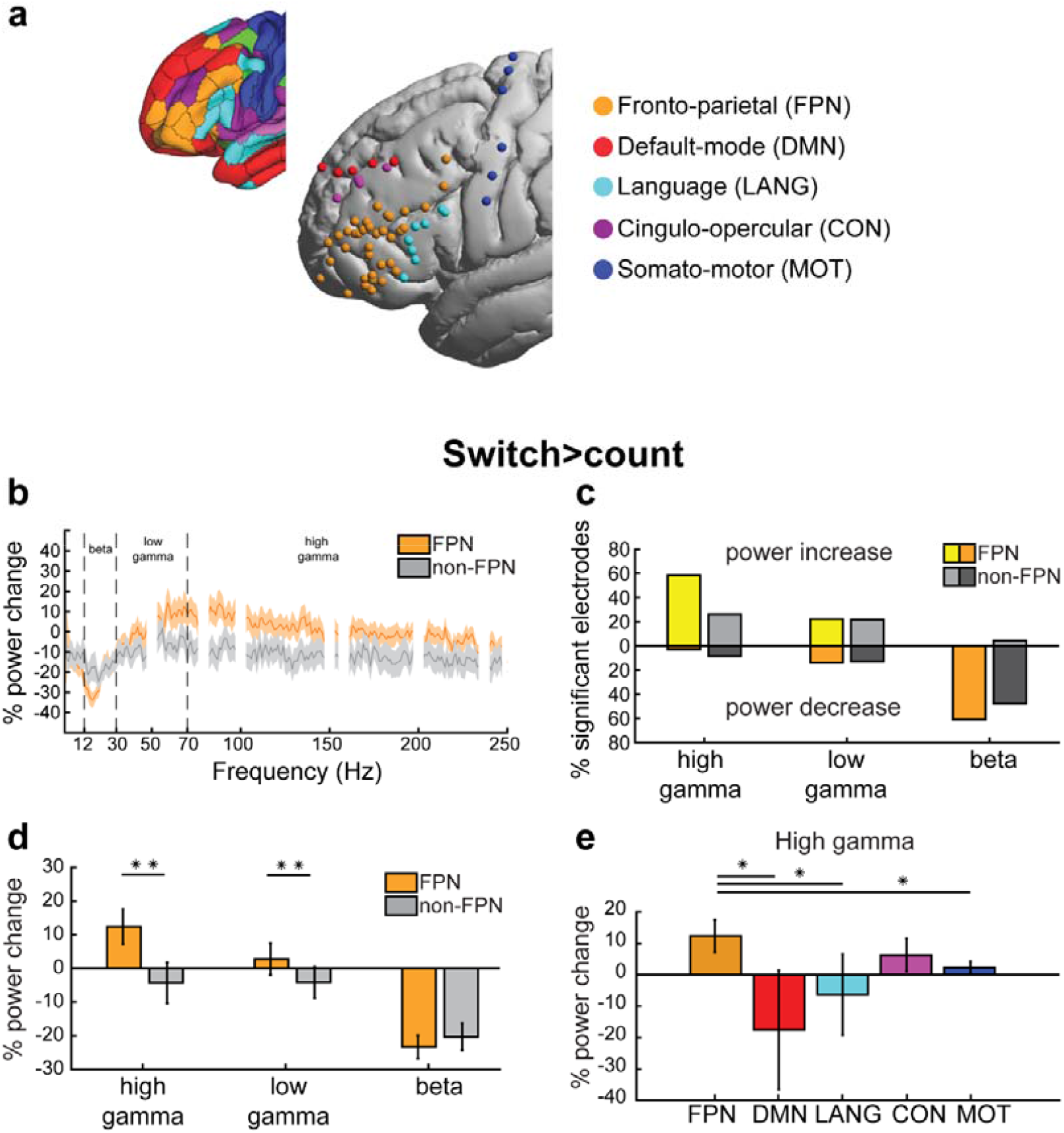
(a) Electrodes colored based on the network label of their nearest vertex. Electrodes on the right hemisphere have been projected onto the left. *Top left inset*: A cortical surface visualization of the 12-network HCP fMRI network parcellation (Ji et al., 2019). (b) PSDs of switch>count contrast averaged across FPN and non-FPN electrodes separately. Notch filtered noise frequency bands (see **Materials and Methods**) are hidden. Shaded areas are SEMs. (c) Percentage of electrodes showing significant power modulations out of all electrodes located within (yellow and orange) and outside (light and dark grey) the FPN. Lighter colored bars (above the zero line) show percentage of electrodes with power increases. Darker colored bars (below the zero line) show percentage of electrodes with power decreases. (d) Average power modulations of all electrodes within and outside the FPN for each frequency band for the switch>count contrast. (e) Average power modulations of all electrodes within each network for the switch>count contrast. Error bars denote SEM. \*\**p*=0.01, \**p*<0.05 one-tailed (linear mixed effects).

First we compared the probability of finding significant electrodes within vs. outside of the FPN network. Of the FPN electrodes, 58.3% showed a significant HG increase in the switch>count contrast, compared to 26.1% of non-FPN electrodes (**Figure 3c**). In contrast, the proportions of electrodes showing LG power increases or beta power decreases were similar within and outside the FPN (LG: 22.2% vs. 21.7%, beta: 61.1% vs. 47.8%). All findings replicated after excluding the motor electrodes (HG: 58.3% vs. 35.3%; LG: 22.2% vs. 29.4%; Beta: 61.1% vs. 47.1%). Therefore, only HG increases, but not LG increases nor beta decreases, were more likely within the FPN compared to outside the network.

**Figure 3d** shows mean percentage power changes in FPN and non-FPN electrodes. HG and LG power increases were significantly stronger in FPN than in non-FPN electrodes [linear mixed effects model (see **Materials and Methods**), HG: t_57_=2.4, LG: t_57_=2.3, both *p*s*=*0.01; p-values are one-tailed due to our directional prediction of gamma increases within the FPN]. There was no significant difference between FPN and non-FPN electrodes for beta power decreases (beta: t_57_=0.3). All findings, except for LG, replicated after excluding the motor electrodes (HG: t_51_=1.9, *p=*0.03; LG: t_51_=1.3, *p*=0.1; beta: t_51_=-0.29). These results highlight that HG power during a demanding task shows localized increases specific to frontal regions of the FPN, while beta decreases were widespread.

We further probed whether HG power increases can distinguish between the finer-grained networks (**Figure 3e**). FPN showed significantly stronger HG increases than all networks (DMN t_54_=1.8; MOT t_54_=1.7; LANG t_54_=1.7; all *p*s*=*0.04 one-tailed, uncorrected) except for CON (t_54_=0.8, *p*=0.2). These results suggest that control-related HG power increases can distinguish the FPN from several of its surrounding networks.

For completeness, we also examined whether the delta, theta or alpha frequency bands distinguished the FPN electrodes from the remainder. We found no evidence for this (maximum t-value 1.5 for alpha, p=0.13 two-tailed), confirming selectivity of our findings to the HG frequency band.

To ensure the robustness of our results, we repeated the analysis using a second canonical fMRI-based cortical parcellation [(Yeo et al., 2011); **see Materials and Methods**]. This parcellation produced 47 FPN electrodes vs 32 non-FPN electrodes. For HG, replicating our previous findings, significant increases were more common in FPN than in non-FPN (52.8% vs. 34.8%). To a lesser extent, the same held for LG (25% vs. 17.4%). Beta decreases were common for both FPN and non-FPN (52.8% vs. 60.9%). Further, HG and LG power increases were significantly stronger in FPN than in non-FPN electrodes (linear mixed effects, HG: t_57_=1.7, *p=*0.04; LG: t_57_=2.4, *p=*0.009; one-tailed) though not after excluding the motor electrodes (HG: t_51_=1.3, *p=*0.1; LG: t_51_=1.6, *p=*0.06; one-tailed). There was a significant difference between FPN and non-FPN electrodes for beta power decreases (beta: t_57_=2.1; *p*=0.02) that did not survive after excluding motor electrodes t_51_=1.6; *p*=0.05). Thus, division using the Yeo et al. FPN parcellation demonstrates similar but statistically weaker results, likely due to its spatially coarser definition of the FPN.

### Count>rest contrast fails to identify the FPN

During the experiment, we had a third experimental condition where patients did not perform any task (rest). To assess the importance of our targeted increased demand manipulation in the switch>count contrast, where the conditions are matched in multiple aspects (e.g. speech, counting), we compared the results with a less controlled count>rest contrast.

The distribution of HG, LG and beta changes in the count>rest contrast is shown in **Figure 4a**. In comparison with switch>count (**Figure 2a**), the count>rest showed a different pattern of gamma power increases, with an apparently more posterior focus. For count>rest, the percentage of significant electrodes was similar between FPN and non-FPN electrodes in all three spectral bands (**Figure 4b**, HG increases: 36.1% vs. 35.3%; LG increases: 27.8% vs. 11.8%; beta decreases: 11.1% vs. 11.8%). The average power modulations did not differ between FPN and non-FPN electrodes for HG (**Figure 4c;** linear mixed effects; with motor electrodes HG: t_57_=0.2 *p*=0.4 one-tailed; without motor electrodes HG: t_57_=0.1 *p*=0.5) but showed an effect for LG (t_57_=1.8 *p*=0.04) that did not survive excluding motor electrodes (LG: t_51_=1.6 *p*=0.06 one-tailed). However, FPN electrodes showed significantly stronger beta power increases (t_57_=2.7 *p*=0.004, without motor electrodes t_51_=2.1 *p*=0.02).

**Figure 4.**
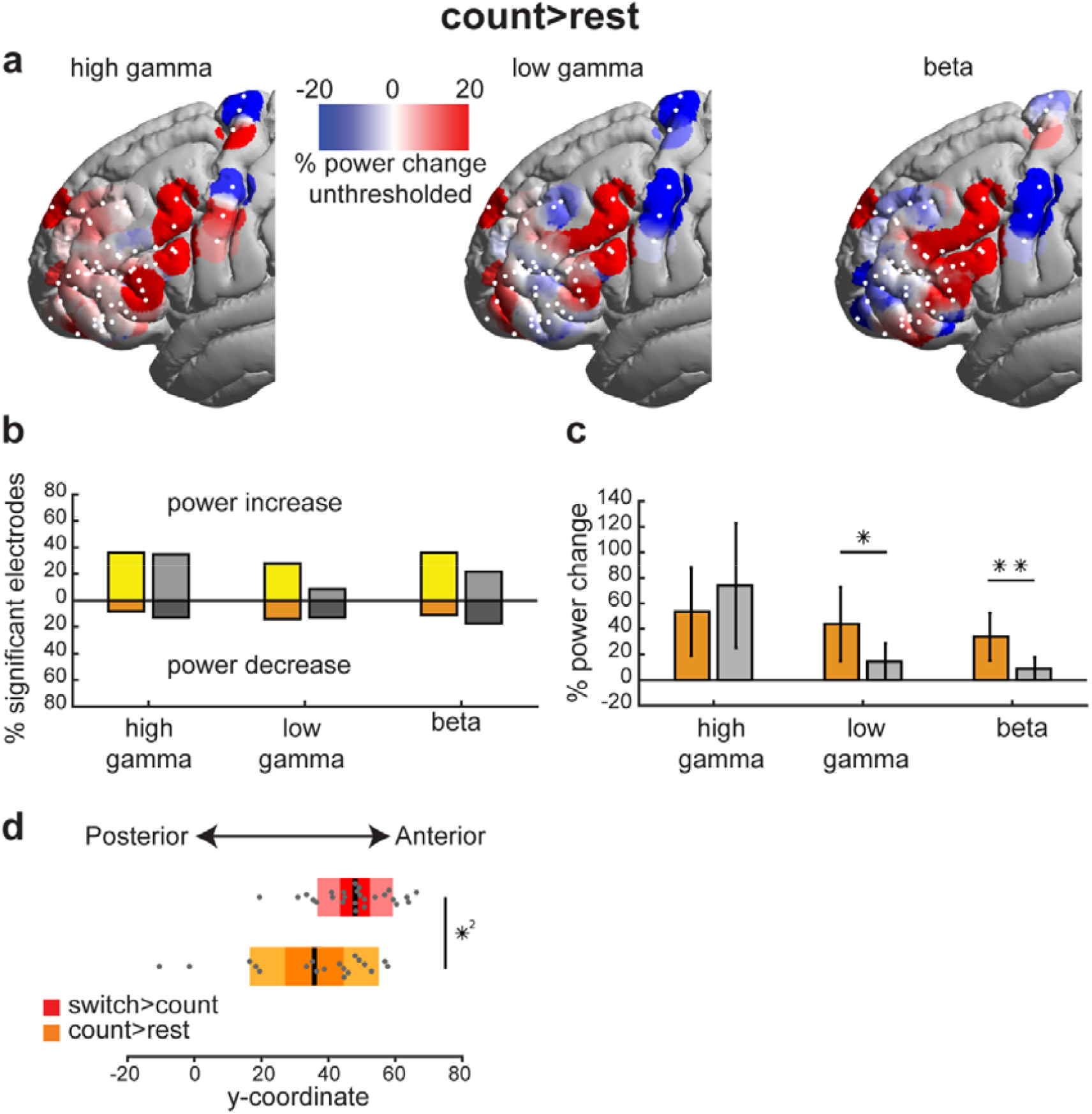
Count>rest spectral power modulations. (a) Unthresholded average smoothed data. Power for each electrode (white dots; including electrodes with non-significant power changes) was spatially smoothed such that the value at each surface vertex is the average of the overlapping powers within a sphere of 10 mm radius. (b) Percentage of electrodes showing significant power modulations out of all electrodes located within (yellow and orange) and outside (light and dark grey) the FPN. Lighter colored bars (above the zero line) show percentage of electrodes with power increases. Darker colored bars (below the zero line) show percentage of electrodes with power decreases. (c) Average power modulations of all electrodes within and outside the FPN for each frequency band for the count>rest contrast. Error bars denote SEM. (d) Box plots of y-coordinate of electrodes with significant HG power increases. Middle black line: mean; darker box limits: 1 SD; lighter box limits: 95% CI. \*\**p*<0.01, \**p*<0.05 one-tailed (linear mixed effects). *^2^*p*<0.05 (Wilcoxon signed rank test)

As suggested by **Figure 4a**, count>rest gamma increases were substantially shifted posteriorly to switch>count increases. To quantify this, we compared y-coordinates (anterior-posterior axis) of electrodes showing significant gamma increases between the two contrasts. The results confirmed a posterior shift for the group of significant HG electrodes in the count>rest contrast (**Figure 4d;** Wilcoxon signed-rank test, HG *p=*0.02) and a similar statistical trend for the LG (*p*=0.07).

Altogether, these results suggest that HG increases can best distinguish frontal control regions in a well-controlled contrast of demanding versus simple cognitive tasks, in line with recent fMRI findings (Assem et al., 2020). The different pattern of HG activity in the less controlled count>rest contrast adds further support to the spatial specificity of findings for a contrast specifically targeting executive control.

## Discussion

We recorded LFP signals using ECoG from the lateral frontal surface of human patients undergoing awake craniotomies for tumor resection. We used a canonical demand on executive control, a contrast of cognitive switching versus simple counting, similar to many manipulations known to recruit FPN regions in fMRI studies (Fedorenko et al., 2013; Shashidhara et al., 2019, 2020, Assem et al., 2020, 2022). The results revealed a circumscribed frontal region that shows increases in HG power. Using two independent fMRI-based frontal parcellations, we found that electrodes overlapping with the FPN showed stronger HG increases that distinguished them from surrounding networks. Our ECoG results thus converge with fMRI in showing fine-grained functional parcellation of cognitive control regions in the lateral frontal cortex.

While many studies using cognitive control tasks have observed increases in gamma power in the frontal lobe, few have attempted to relate HG topographies to fMRI-based frontal lobe parcellation. One large-scale ECoG study used resting-state LFP and found that grouping electrodes based on the synchrony of their time-series in the delta band (1-4 Hz) mapped to some extent to the canonical functional divisions in fMRI (Betzel et al., 2019). Here we show that high gamma power during a cognitive control task can reliably distinguish the FPN from other frontal networks. Gamma increases are generally interpreted to reflect localized task processing. A number of studies link gamma to fMRI activations in early cortical regions (Nir et al., 2007; Engell et al., 2012; Hermes et al., 2012). The current results extend these reports to the domain of cognitive control in frontal regions.

Because HG and not LG reliably distinguished frontal control regions, our results support previous suggestions for different physiological origins of high and low gamma (Crone et al., 2006; Buzsáki and Wang, 2012; Lachaux et al., 2012). In contrast to focal increases in the gamma range, we observed spatially broad decreases in power in the beta band (and other lower frequencies) which were not confined to the FPN. It has been previously shown that synchronization in lower frequency bands between fronto-parietal regions increased during executive tasks e.g. (Voytek et al., 2015). Combined with our findings, this suggest that increases in beta synchrony may be accompanied by decreases in power. Concerning the relation between gamma increases and beta decreases, one suggestion is that these reflect two sides of the same process, a rotated power spectrum around a middle range frequency (Podvalny et al., 2015; Helfrich and Knight, 2016). Recent evidence, however, argues against this simple interpretation, showing that depending on the cortical region, increases in high gamma power are not necessarily accompanied by decreases in low frequency power (Fellner et al., 2019). In line with this, the current results also showed that beta decreases were more spatially broad and were not necessarily accompanied by gamma increases. Another framework proposes a hybrid spiking-synaptic plasticity working memory model, in which bursts of spikes (gamma increases) in superficial layers serve to encode and maintain working memory content, while beta, which is assumed to have an inhibitory role, is suppressed in deeper layers to allow superficial gamma bursts (Lundqvist et al., 2011; Miller et al., 2018). This model is specific for regions that are involved in working memory processes. As it stands, however, this model may need extension to explain beta power decreases within but also outside of FPN.

For the count>rest contrast, gamma modulations were not found selectively within the FPN. While the switch>count contrast compared conditions that are closely matched on features such as speech and counting task, the count>rest contrast is less controlled. Accordingly, changes in activity may be related to several differences between the conditions, most prominently speech production since the patient remained silent during the rest condition. Plausibly, posterior gamma increases might reflect articulation or language related processing in the posterior inferior frontal gyrus (Basilakos et al., 2018).

Our findings open the door for extending clinical functional mapping to the domain of cognitive/executive control during tumor resection surgeries. Damage to control regions is associated with disorganized behaviour (Glascher et al., 2010, 2012, Woolgar et al., 2010, 2018; Warren et al., 2014) and poorer recovery following neurosurgery (Romero-Garcia et al., 2019). During surgeries, electrical stimulation is commonly used to map motor and language functions, where activity is confined to well-defined areas with clear behavioural responses, so that resection of eloquent tissue can be avoided. In higher association areas and in particular for executive control regions, the use of stimulation is more challenging, primarily because of the complex nature of the mapped functions and the distributed areas that support them. In addition, intraoperative stimulation is time-consuming while neurosurgeons search for the areas responsive to the tested functions with increased risk of seizures. Current mapping approaches to assess executive regions intraoperatively are limited, with only sparse evidence where direct electrical stimulation was used (Wager et al., 2013; Puglisi et al., 2018, 2019; Mandonnet et al., 2020). ECoG has the potential to provide complementary information to guide stimulation and clinical decision making, especially for executive control functions. The link between increased gamma power and the FPN with increased cognitive demand establishes a critical anatomo-functional foundation for this approach. Matching the fine scale of regional specializations within the frontal cortex, our results suggest that distinct functional regions may be practically mapped in the context of awake tumor surgery.

## Data Availability

Due to clinical ethical considerations and in accordance with the ethics approval for the study by the East of England - Cambridge Central Research Ethics Committee (REC reference 16/EE/0151), the data cannot be shared or become publicly available.

## Acknowledgments

The Royal Society provided financial support in the form of a Royal Society Dorothy Hodgkin Research Fellowship to YE (DH130100). Cambridge Commonwealth European and International Trust provided financial support in the form of a Yousef Jameel scholarship to MA. Guarantors of Brain provided financial support in the form of a Post-Doctoral Fellowship award to RRG. The National Institute for Health Research (NIHR, UK) provided financial support in the form of a Clinician Scientist Award 35 to SJP (ref: NIHR/CS/009/011). The Brain Tumour Charity provided financial support in the form of a grant award to MGH (ref: RG86218). J.D was funded by a Medical Research Council grant (MC_UU_00005/6). This work was supported by the NIHR Cambridge Biomedical Research Centre (BRC-1215-20014) and NIHR Applied Research Centre. All the sponsors had no role in the design or conduct of this research. We thank Mallory Owen for help with administering and analyzing neuropsychological tests.

## TOP related statements

- Due to clinical ethical considerations and in accordance with the ethics approval for the study by the East of England - Cambridge Central Research Ethics Committee (REC reference 16/EE/0151), the data cannot be shared or become publicly available.
- No part of the study procedures or analysis was pre-registered prior to the research being conducted.
- We report how we determined our sample size, all data exclusions, all inclusion/exclusion criteria, whether inclusion/exclusion criteria were established prior to data analysis, all manipulations, and all measures in the study.

## Open Access statement

For the purpose of open access, the author has applied a Creative Commons Attribution (CC BY) license to any Author Accepted Manuscript version arising from this submission.

